# Estimates of Cross-Border Menthol Cigarette Sales Following the Comprehensive Tobacco Flavor Ban in Massachusetts

**DOI:** 10.1101/2022.04.24.22274236

**Authors:** Jacob James Rich

## Abstract

On June 1, 2020, Massachusetts became the first state in the US to ban all flavored tobacco product sales, including menthol cigarettes. Recent research has estimated the reduction in cigarette sales in Massachusetts following the comprehensive tobacco flavor ban, but noted that missing data on border states was a major limitation of the findings. This letter replicates the procedures of Asare et al. with 1540 state-months and then adds Asare et al.’s missing states with 2420 total observations for the period January 2017 to July 2021. The replication confirms Asare et al.’s adjusted estimate for the reduction in menthol cigarettes, which falls within their 95% confidence interval. However, assigning Massachusetts and its bordering states as a single treatment group leads to an increase of 191.95 (95% CI, 96.82 to 287.09) total cigarette packs sold per 1000 people in the six-state region. In the 12-month period following the comprehensive flavor ban in Massachusetts, the state sold 29.96 million fewer cigarette packs compared to the prior period. However, a total of 33.36 million additional cigarette packs were sold during the same post-ban period in the counties that bordered Massachusetts. Given decreasing rates of smoking in all five bordering states between 2019 and 2020, the increase in border-state cigarette sales following the comprehensive flavor ban should be interpreted as a lower-bound estimate for cigarettes that were ultimately consumed in Massachusetts.

## Methods

Using monthly MSAi retailer to wholesaler cigarette shipment data, Asare et al.’s difference-in-differences estimates are replicated to measure changes in state-level sales of menthol, nonflavored, and total cigarette packs. These estimates are then compared to an identical specification that adds Washington, DC and the 15 missing states without tobacco flavor bans during the same period. Massachusetts and its bordering states are then evaluated as treatment groups.

Descriptive statistics for the variables of interest matched the means in the original analysis, minus those for cigarette prices, household income, and COVID-19 case rates (Supplement).^[2]^ There are also outcome differences because the Nielsen Retail Scanner Data measuring cigarette sales in the original letter only represent approximately 30% of all US mass merchandiser sales volume.^[3]^ In contrast, the MSAi data represent all cigarette distribution in the US. In order to match Asare et al.’s estimates, survey weights were removed from most of the IPUMS-CPS data.^[4]^ All means that could not be matched were then verified by reports from the data-collecting agencies.

## Results

The procedures of Asare et al. are replicated with 1540 state-months and then expanded with 2420 total observations for the period January 2017 to July 2021 (Supplement), with nondivergent trends in cigarette sales before treatment. After the prohibition, the replication confirms Asare et al.’s adjusted estimate for menthol cigarettes, which falls within their 95% confidence interval (Table 1). The complete model then estimates that the monthly sales of cigarette packs per 1000 people in Massachusetts decreased for menthol 292.66 (95% CI, -557.17 to -28.15) and increased for nonflavored 224.16 (95% CI, -144.30 to 592.64), resulting in a total reduction of 68.49 (95% CI, -608.36 to 471.37). However, assigning the border states as a single treatment group then leads to an increase of 80.57 (95% CI, 38.81 to 122.34) menthol and 119.88 (95% CI, 50.44 to 189.32) nonmenthol cigarette packs sold per 1000 people in the bordering states. Finally, assigning Massachusetts and its bordering states as a single treatment group leads to an increase of 191.95 (95% CI, 96.82 to 287.09) total cigarette packs sold per 1000 people in the six-state region.

**Table 1.** Difference-in-Differences Estimates for Cigarettes Sold per 1000 Population.

| Variable | Asare.et.al. | 95% CI<br>(Low High) |  | Replication | 95% CI<br>(Low High) |  | Complete<br>Model | 95% CI<br>(Low High) |  | Border<br>Model | 95% CI<br>(Low High) |  | Regional<br>Model | 95% CI<br>(Low High) |  |
| --- | --- | --- | --- | --- | --- | --- | --- | --- | --- | --- | --- | --- | --- | --- | --- |
| Menthol | *** -372.27 | -428.9 | -315.64 | * -245.66 | -435.20 | -56.12 | * -292.66 | -557.17 | -28.15 | *** 80.57 | 38.81 | 122.34 | *** 72.57 | 34.69 | 110.44 |
| Nonmenthol | *** 120.25 | 72.61 | 167.88 | 243.35 | -54.13 | 540.83 | 224.16 | -144.30 | 592.64 | ** 119.88 | 50.44 | 189.32 | **119.38 | 50.92 | 187.85 |
| All Cigarettes | *** -282.65 | -356.07 | -209.23 | -2.31 | -418.54 | 413.92 | -68.49 | -608.36 | 471.37 | *** 200.46 | 103.21 | 297.70 | ***191.95 | 96.82 | 287.09 |
| Observations | 1652 |  |  | 1540 |  |  | 2420 |  |  | 2260 |  |  | 2205 |  |  |

Note: \* $p < .05$ , \*\* $p < .01$ , \*\*\* $p < .001$

In the 12-month period following the comprehensive flavor ban in Massachusetts, the state sold 29.96 million fewer cigarette packs compared to the prior period. However, a total of 33.36 million additional cigarette packs were sold during the same post-ban period in the counties that bordered Massachusetts (Figure 1). Considering the change in sales for the entire six-state region, there was a net increase of 7.21 million additional cigarette packs sold following the Massachusetts comprehensive flavor ban, a 1.27% increase from the prior 12-month period.

**Figure 1.**
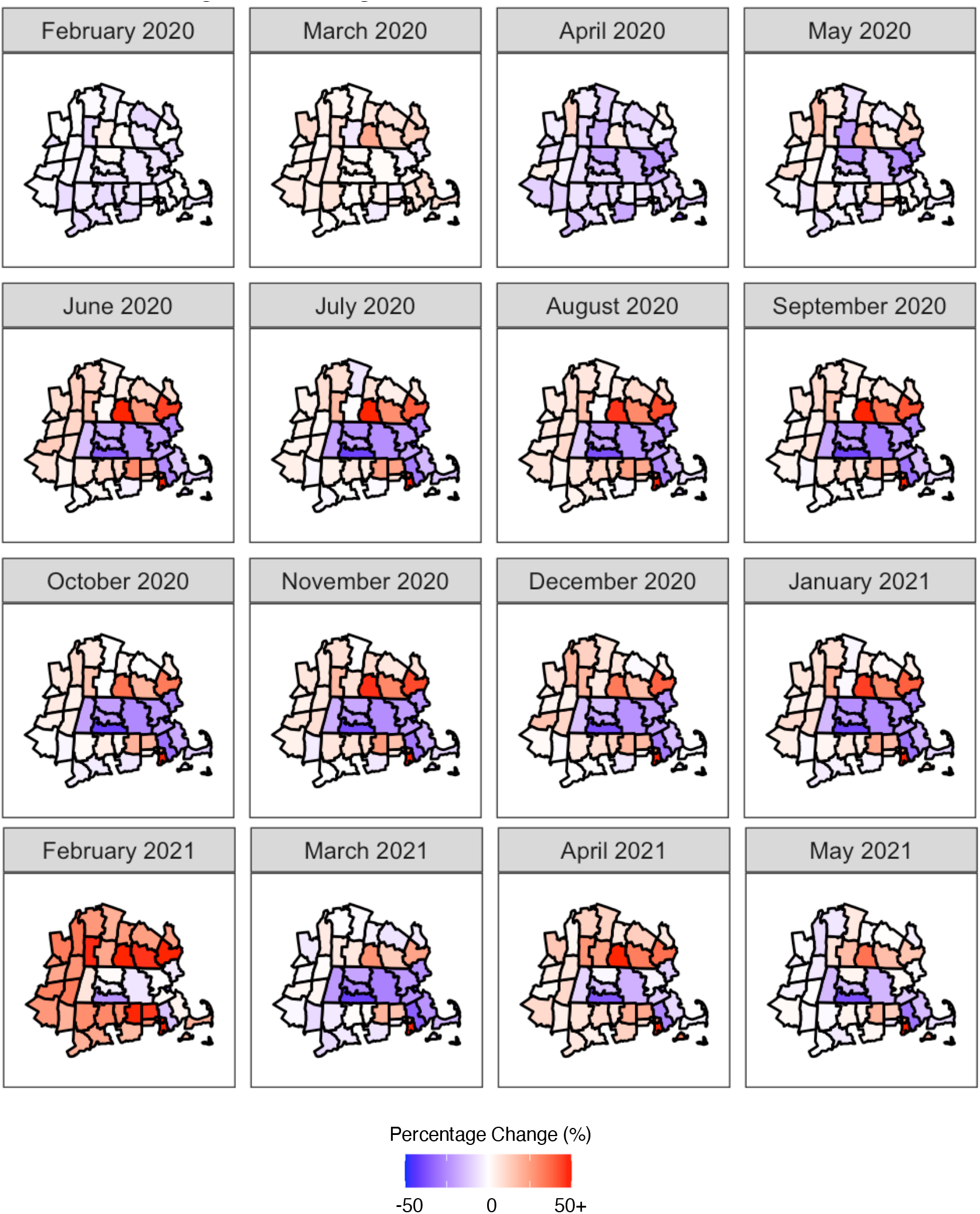
Percent Change in Total Cigarette Pack Sales from Previous Year. Note: Percentage change references the same month in the previous year.

## Discussion

State-level prohibitions on flavored tobacco sales are far less effective when bordering states and counties provide access to prohibited products. Additionally, tobacco flavor bans may lead to net increases in tobacco sales when outside localities charge lower excise taxes. Given decreasing rates of smoking in all five bordering states between 2019 and 2020,^[5]^ the increase in border-state cigarette sales following the comprehensive flavor ban should be interpreted as a lower-bound estimate for cigarettes that were ultimately consumed in Massachusetts.

## Supporting information

Supplemental Description

Predictor Variable Data

## Data Availability

All predictor variables in the present work are available in the supplements. Most of the outcome data will be released by the CDC at a later date.

