## Supplemental Description for "Estimates of Cross-Border Menthol Cigarette Sales Following the Comprehensive Tobacco Flavor Ban in Massachusetts"

**eMethods 1.** Data Collection and Construction
**eMethods 2.** Difference-in-Differences Specification
**eFigure 1.** Percent Change in Menthol Cigarette Pack Sales from Previous Year
**eTable 1.** Comparative Descriptive Statistics for January 2019 to July 2021
**eTable 2.** List of States in Each Specification
**eReferences**

This supplementary material has been provided by the author to give readers additional information about his work.

**eMethods 1.** Data Collection and Construction

The predictor variables in the difference-in-differences analysis were obtained or constructed from public sources and are available on *medRxiv* for replication purposes. The cigarette distribution outcome data are not provided, but total cigarette sales volume is collected by Orzechowski and Walker and will be released by the Centers for Disease Control and Prevention (CDC) in a later version of the Tax Burden on Tobacco annual compendium.[1]

eTable 1. displays the proportions for the predictor variables published by Asare et al. and collected for this replication. Household weights are applied to sex and income, but all other CPS variables are left unweighted to match Asare et al.'s estimates.

Wholesaler shipment data are an almost-perfect proxy for monthly sales volume, but they contain no information on the final price sold at the register. There are no public monthly data on cigarette price fluctuations like used by Asare et al. Public data from the Tax Burden on Tobacco contain average annual cigarette pack price estimates, but they currently only describe years up to 2019.[1] However, the US Census Bureau collects tobacco excise tax revenue at the state & local levels in its Quarterly Summary of State & Local Tax Revenue Data.[2] According to Table 7 of the Statistical Abstract of North Carolina Taxes 2020, 88.1% of state & local tobacco tax collections come from cigarettes.[3] Since federal taxes are equal state-to-state and subtracting state & local taxes from the final prices yields average pre-tax cigarette prices within $1.00 (except Alaska, Hawaii, Massachusetts, and New York), the correlation between final prices and state & local taxes for cigarettes is $r = 0.963$.[3] Since 96.3% of the final price fluctuation is captured in the state & local taxes, we use them as a proxy for register prices, which are calculated by dividing the Census quarterly estimates into three equal monthly estimates, multiplying them by 0.881, and dividing by the total number of cigarettes shipped by wholesalers that month.

Unlike the prices used in Asare et al., these tax estimates are not specific to the cigarette flavor represented by the outcome variable. However, when menthol cigarettes were banned in Massachusetts, it would be incorrect to code the menthol cigarette price at $0, when in reality the price would equal ∞. What Asare et al. did in this circumstance is not described.

COVID-19 case numbers published by Asare et al. also do not match the replicated estimates in eTable 1. Asare et al. sourced their case numbers from the New York Times while this replication sourced from the CDC. However, the mean COVID-19 case numbers per 1000 population published by Asare et al. could not possibly reflect the entire period. Assuming the authors only meant to estimate cases during the pandemic, the replicated mean COVID-19 case numbers only reflect April 2020 to July 2021.[4]

**eMethods 2.** Difference-in-Differences Specification

$$Y_{st} = \alpha_0 + \alpha_1 \mathrm{DiD}_{st} + \alpha_2 \mathrm{Tr}_{st} + \alpha_3 \mathrm{Ti}_{st} + \alpha_4 P_{st} + \beta \mathrm{X}_{st} + \beta \gamma_{st} + \xi_{st}, \; s = \text{state} \;\&\; t = \text{time}$$

where $Y_{st}$ is the monthly sales of menthol, non-menthol, and all cigarettes per 1000 people; $\alpha_0$ the constant; $P_{st}$ the annual average state & local tax (price proxy) of a cigarette pack; DiD the difference-in-differences estimator for the Massachusetts flavor ban, which is an interaction between the Tr and Ti treatment-period dummy variables; X a vector of the variables listed in eTable 1. (dropping the last of each category to set it as the reference variable); $\gamma_{st}$ a vector of state, month, and year fixed effects; and $\xi_{st}$ the error.

Following the comprehensive tobacco flavor ban in Massachusetts, some counties reported negative menthol cigarette shipments, which represent returns. We reduced the previous month's shipment estimates for those counties by those magnitudes.

**eFigure 1.** Percent Change in Menthol Cigarette Pack Sales from Previous Year

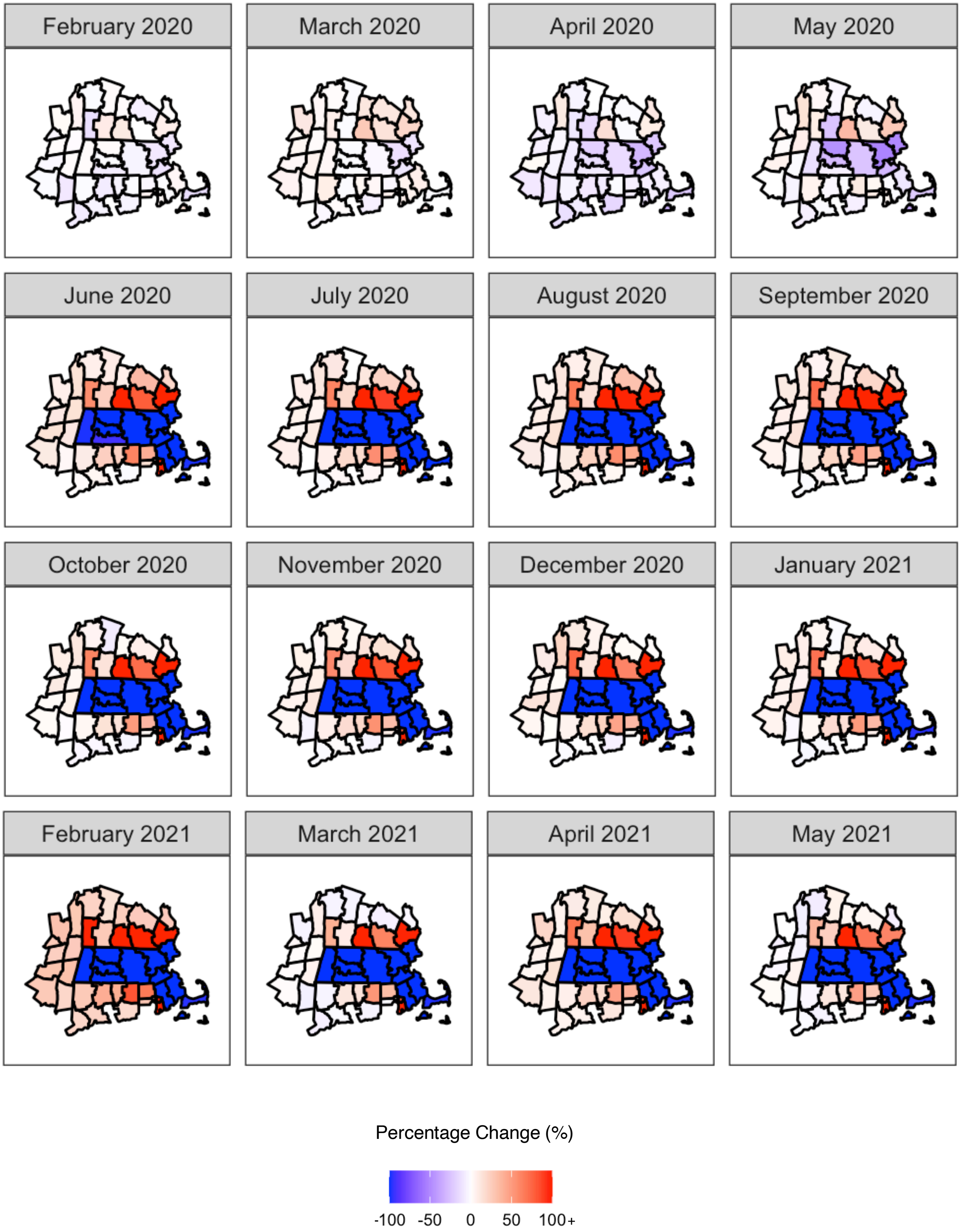


Note: Percentage change references the same month in the previous year.

**eTable 1.** Comparative Descriptive Statistics for January 2019 to July 2021

| Variable | Asare et al. MA | MA Replication | Asare et al. Comparison States | Comparison Replication | Complete Model | Complete Model no MA |
|---|---|---|---|---|---|---|
| Menthol Sales (/1000 p.) | N/A | 444.19 | N/A | 1197.88 | 484.48 | 484.95 |
| Nonmenthol Sales | N/A | 1183.85 | N/A | 1967.34 | 823.59 | 819.31 |
| All Cigarettes Sales | N/A | 1628.04 | N/A | 3165.22 | 1308.07 | 1304.27 |
| Menthol Price (average) | 10.16 | N/A | 6.79 | N/A | N/A | N/A |
| Nonmenthol Price | 9.87 | N/A | 6.58 | N/A | N/A | N/A |
| All Cigarettes Price | 9.94 | N/A | 6.65 | N/A | N/A | N/A |
| All Cigarettes Tax | N/A | 3.46 | N/A | 1.52 | 1.19 | 1.16 |
| Male | 48.26 | 48.26 | 48.40 | 48.53 | 48.54 | 48.55 |
| Female | 51.74 | 51.74 | 51.60 | 51.47 | 51.46 | 51.45 |
| Married | 49.28 | 49.32 | 52.24 | 52.12 | 52.00 | 52.07 |
| Unmarried | 50.72 | 50.68 | 47.76 | 47.88 | 48.00 | 47.93 |
| Age <25 | 28.06 | 28.11 | 29.70 | 29.87 | 29.84 | 29.89 |
| 25-44 | 26.68 | 26.70 | 24.61 | 24.70 | 24.95 | 24.90 |
| 45-64 | 27.52 | 27.53 | 26.85 | 26.77 | 26.49 | 26.46 |
| Over 64 | 17.74 | 17.67 | 18.84 | 18.66 | 18.72 | 18.75 |
| Asian | 7.20 | 7.17 | 3.83 | 3.86 | 4.40 | 4.33 |
| Black | 7.23 | 7.17 | 13.51 | 13.89 | 11.44 | 11.55 |
| White | 85.57 | 82.70 | 79.24 | 79.07 | 80.01 | 79.95 |
| Other | 3.00 | 2.96 | 3.41 | 3.19 | 4.14 | 4.17 |
| No high school | 10.60 | 10.65 | 13.87 | 14.22 | 13.47 | 13.54 |
| High school | 24.09 | 24.17 | 29.15 | 29.12 | 28.82 | 28.94 |
| Some college | 21.08 | 21.11 | 26.80 | 26.68 | 26.60 | 26.74 |
| College | 44.23 | 44.07 | 30.18 | 29.98 | 31.11 | 30.77 |
| Income <$10 000 | 19.03 | 4.64 | 19.49 | 4.87 | 4.87 | 4.88 |
| $10 000-$29 999 | 10.12 | 12.04 | 14.31 | 16.92 | 16.63 | 16.76 |
| $30 000-$59 999 | 15.69 | 18.67 | 22.71 | 27.13 | 26.67 | 26.89 |
| $60 000-$149 000 | 32.71 | 38.57 | 32.54 | 37.92 | 38.07 | 38.05 |
| $150 000+ | 22.46 | 26.09 | 10.95 | 13.16 | 13.77 | 13.43 |
| Unemployment | 4.97 | 4.87 | 4.92 | 4.71 | 4.62 | 4.62 |
| COVID-19* | 13.84 | 6.45 | 14.13 | 6.58 | 2.93 | 2.89 |
| Observations | 59 | 55 | 1593 | 1485 | ** 2475 | 2420 |

* Period April 2020 to July 2021

** Includes NYC, which is not included in the “Complete Model” regression.

**eTable 2.** List of States in Each Specification

| State FIPS | State | Asare et al. | Surrounding States | Complete Model |
|---|---|---|---|---|
| 1 | Alabama | 1 | 0 | 1 |
| 2 | Alaska | 0 | 0 | 1 |
| 4 | Arizona | 1 | 0 | 1 |
| 5 | Arkansas | 1 | 0 | 1 |
| 6 | California | 0 | 0 | 0 |
| 8 | Colorado | 0 | 0 | 0 |
| 9 | Connecticut | 1 | 1 | 1 |
| 10 | Delaware | 0 | 0 | 1 |
| 11 | District of Columbia | 0 | 0 | 1 |
| 12 | Florida | 1 | 0 | 1 |
| 13 | Georgia | 1 | 0 | 1 |
| 15 | Hawaii | 0 | 0 | 1 |
| 16 | Idaho | 0 | 0 | 1 |
| 17 | Illinois | 0 | 0 | 0 |
| 18 | Indiana | 1 | 0 | 1 |
| 19 | Iowa | 0 | 0 | 1 |
| 20 | Kansas | 1 | 0 | 1 |
| 21 | Kentucky | 1 | 0 | 1 |
| 22 | Louisiana | 1 | 0 | 1 |
| 23 | Maine | 0 | 0 | 0 |
| 24 | Maryland | 1 | 0 | 1 |
| 25 | Massachusetts | 1 | 0 | 1 |
| 26 | Michigan | 1 | 0 | 1 |
| 27 | Minnesota | 0 | 0 | 0 |
| 28 | Mississippi | 1 | 0 | 1 |
| 29 | Missouri | 1 | 0 | 1 |
| 30 | Montana | 0 | 0 | 1 |
| 31 | Nebraska | 0 | 0 | 1 |
| 32 | Nevada | 1 | 0 | 1 |
| 33 | New Hampshire | 0 | 1 | 1 |
| 34 | New Jersey | 1 | 0 | 1 |
| 35 | New Mexico | 0 | 0 | 1 |
| 36 | New York | 0 | 1 | 0 |
| 37 | North Carolina | 1 | 0 | 1 |
| 38 | North Dakota | 0 | 0 | 1 |
| 39 | Ohio | 1 | 0 | 1 |
| 40 | Oklahoma | 1 | 0 | 1 |
| 41 | Oregon | 1 | 0 | 0 |
| 42 | Pennsylvania | 1 | 0 | 1 |
| 44 | Rhode Island | 0 | 1 | 1 |
| 45 | South Carolina | 1 | 0 | 1 |
| 46 | South Dakota | 0 | 0 | 1 |
| 47 | Tennessee | 1 | 0 | 1 |
| 48 | Texas | 1 | 0 | 1 |
| 49 | Utah | 0 | 0 | 1 |
| 50 | Vermont | 0 | 1 | 1 |
| 51 | Virginia | 1 | 0 | 1 |
| 53 | Washington | 1 | 0 | 1 |
| 54 | West Virginia | 0 | 0 | 1 |
| 55 | Wisconsin | 1 | 0 | 1 |
| 56 | Wyoming | 0 | 0 | 1 |

[2] US Census Bureau, "Quarterly Summary of State & Local Tax Revenue Data Tools & Apps," US Department of Commerce, October 8, 2021.

[3] North Carolina Department of Revenue, "Statistical Abstract of North Carolina Taxes 2019," North Carolina Department of Revenue, Raleigh, NC, July 13, 2021.

[4] CDC Case Task Force, "United States COVID-19 Cases and Deaths by State over Time," Centers for Disease Control and Prevention, Atlanta, GA, April 23, 2022.
